# Stroke rates, risk factors, and aspirin prescribing trends in the Canadian Fabry Disease Initiative cohort

**DOI:** 10.1101/2024.07.17.24310571

**Authors:** Emilie T. Théberge, Caroline Selvage, Anita Thomas, Kaye M. LeMoine, Rebecca Robichaud, Lily Zhou, Darwin F. Yeung, Michael L. West, the CFDI investigators group, Sandra Sirrs, Anna Lehman

## Abstract

**Background:** Fabry disease (FD) is an X-linked disorder caused by deleterious variants in *GLA.* Cardiovascular disease (CVD) causes premature mortality in FD. Hope for aspirin (acetylsalicylic acid, ASA) to reduce CVD risks in FD as primary prevention may have been tempered by the 2018 ARRIVE, ASCEND, and ASPREE clinical trials. It is unclear how new ASA guidance applies to FD patients, who have a high rate of young-onset, small vessel stroke compared with the general population.

**Methods:** Longitudinal data spanning 2007-2023 from patients in the Canadian Fabry Disease Initiative (CFDI) were analyzed retrospectively. Incident stroke and transient ischemic attack (TIA), other CVD events, FD-specific risk factors, and ASA/antiplatelet (“ASA/AP”) prescription before and after 2018 were compared between groups who never had an event (“primary prevention group”) to those who had incident stroke/TIA during the study. Stroke/TIA rates were compared between the sexes, by *GLA* variant severity, and between the CFDI to Canadian statistics. 10-year atherosclerotic CVD (ASCVD) risk was calculated using the 2013 ACC/AHA risk calculator. ASA/AP prescription rate was compared before and after 2018.

**Results:** Out of 641 patients, 57 had an incident stroke/TIA during the study, and 193 with complete data remained in the primary prevention group. Stroke/TIA rates were significantly higher among male patients (0.026/year) than females (0.0098/year), and higher among patients with severe *GLA* variants (males: 0.031, females: 0.0096) compared to those with attenuated variants (males: 0.011, females: 0.0088). No patients under 60 years at their incident stroke/TIA had high (≥10%) calculated 10-year ASCVD risk. Fewer patients were prescribed ASA/AP for primary prevention after 2018.

**Conclusions:** There was a high incidence of stroke/TIA in the younger CFDI cohort compared to the general Canadian population, despite low levels of traditional vascular risk factors as represented in 10-year estimated ASCVD risk. Primary prevention use of ASA has declined.

## Introduction

Fabry disease (FD) (OMIM #301500) is a rare X-linked disorder caused by mutations in the galactosidase alpha (*GLA)* gene affecting approximately 1 in 40,000 males and 1 in 20,000 females^1^. Deficient alpha-galactosidase A (a-gal A) enzyme activity results in a buildup of globotriaosylceramide (Gb3) in lysosomes of most cardiac and renal cell types, as well as vascular endothelium^2^. Cardiovascular disease (CVD) is the main cause of death in FD patients^3^.

Stroke is 12 times more frequent in patients with FD than the general population and occurs at significantly younger ages^4,5^. The mean age at first stroke from the American Fabry Registry^5^ patients analyzed between 2001-2007 was 40 years for males and 46 years for females, in contrast to the mean age of first stroke in the general American population of ages 76 and 81, respectively^6^. Ischemic-type strokes and transient ischemic attacks (TIAs) predominate among FD patients (86%)^7^, with the large majority being lacunar (small vessel) or of cryptogenic (unknown) etiology^5,7^.

Aspirin, or acetylsalicylic acid (ASA), is a non-steroidal anti-inflammatory drug that suppresses normal platelet function. It is widely prescribed for both primary and secondary prevention of major adverse cardiovascular events (MACE), including stroke/TIAs, although the actual number of people in Canada who are taking ASA is not known^8^. Atherosclerotic cardiovascular disease (ASCVD) is the most common pathophysiology underlying ischemic stroke, causing luminal narrowing of major blood vessels and/or plaque rupture and thrombus leading to cerebral embolism. Robust evidence has shown benefits of ASA (and other antiplatelet agents) in secondary prevention of CVD^9^; However, recent publications in 2018 from the ASCEND^10^, ARRIVE^11^ and ASPREE^12^ clinical trials have challenged the assumption of ASA’s benefit in primary prevention against ischemic MACE outcomes in more general populations.

In the ARRIVE trial, among men >55 years old and women >60 with “moderate” estimated risk of a first CVD event, authors could not confirm the efficacy of ASA in primary prevention, with mixed results of MACE outcomes and stroke showing conflicting trends of log-rank MACE risk in ASA patients relative to placebo^11^. In the ASCEND trial, ASA use prevented serious vascular events in men and women over 40 years old with diabetes and no evident CVD at trial entry; however, it also caused major bleeding events; authors reported the absolute benefits were largely counterbalanced by the bleeding hazard^10^. Lastly, in the ASPREE trial, low-dose ASA as a primary prevention strategy in older adults (>65 at trial recruitment) resulted in a significantly higher risk of major hemorrhage and did not result in a significantly lower risk of CVD than placebo^12^. As a result of these trials, Canadian stroke prevention guidelines published in 2020 have revised recommendations regarding ASA use to be more individualized, and less favorable for the use of ASA in primary prevention contexts^8^.

Single or dual antiplatelet therapy is common in primary and secondary prevention of ischemic strokes in the general population, but the efficacy in FD patients is unknown. It is uncertain how current ASA recommendations should translate to a population with FD, who have a high *a priori* risk of early-onset strokes/TIAs before the age of 60 years.

The aims of this study are thus to investigate: 1) Incident first stroke/TIA rate in the Canadian Fabry Disease Initiative (CFDI) cohort; 2) ASCVD risk among FD patients with prior stroke/TIA compared to patients without prior MACE; and 3) Antiplatelet prescribing practices in the time periods before and after the 2018 trials, to observe whether or not the publication of these trials may be associated with reduced prescribing.

## Methods

### Cohort characteristics and variables

FD patients were recruited to the CFDI, a prospective national disease registry operating at 19 sites in Canada that began recruitment in 2007. The CFDI registry study is approved by local research ethics boards and all patients provide written informed consent. All data received were de-identified. *GLA* variant class was classified into a binary of “severe” and “attenuated,” using a cut-off of <5% a-gal A enzyme activity as observed in males, to define the former. Details of the variant classification methodology are detailed in the Supplemental Methods.

Demographic, clinical and laboratory values were recorded into the CFDI registry database, powered by the Omda informatics system. Sex was self-reported as male or female; age was derived via year of birth; a-gal A enzyme, globotriaosylsphingosine (lyso-Gb3) and estimated glomerular filtration rate (eGFR) measurements were ascertained via clinical laboratories using blood samples; left ventricular mass index (LVMI) was ascertained by echocardiogram; dyslipidemia, diabetes and hypertension status were ascertained by treating physicians; and smoking status was self-reported by patients. All variables except year of birth and *GLA* variant were time-varying, so data were extracted at timepoints relevant to corresponding analyses.

### MACE and stroke/TIA rate

MACE events were quantified in these analyses as stroke/transient ischemic attack (TIA), myocardial infarction (MI), or percutaneous coronary intervention (PCI)/coronary artery bypass grafting (CABG). Heart failure (HF) events were not quantified for two reasons: 1) From a lack of evidence of ASA aiding in primary prevention of HF in general populations; 2) The majority of the clinical trials and United States Preventive Service Task Force (USPSTF) 2022 guidelines informing conflicting of evidence did not include HF in their composite MACE outcome^10,11,13^, with the exception of the ASPREE trial^12^. Cardiovascular deaths from non-stroke causes were not counted.

Only patients with their first stroke/TIA events occurring during the study period were retained so as to be able to calculate rate per patient years, and patients with their incident stroke/TIA prior to the study were excluded from downstream analyses. The year and occurrence of a prior stroke and/or TIA were analyzed from fields summarizing medical history prior to enrollment and events occurring while in the study. It was not possible to parse out if patients had a stroke versus a TIA, as the input field in the study combined them together, therefore they were analyzed as a combined unit (“stroke/TIA”).

Overall stroke/TIA rate and first stroke/TIA rate per patient year were calculated by sex and *GLA* variant subtype of attenuated or severe. Average number of years in the study per patient were calculated by dividing the total number in the subgroup by the total number of patient years. Subgroups of patient year calculations were performed by sex, *GLA* and age group (<40, 40-59, 60+).

Canadian stroke statistics were assessed to compare rates of stroke/TIA incidence in the CFDI cohort compared to the general population, using sex- and age-stratified data from 2012-2013^14^. Of note, Canadian statistics detail stroke and not TIA, but also include recurrent strokes.

### Lacunar stroke prevalence

Qualitative comments entered by study physicians at the 14 sites of brain magnetic resonance imaging (MRI) and computed tomography (CT) results were available and reviewed for the patients with incident stroke/TIA event occurring while in the study. The following English and French keywords were searched: “lacunar”, “lacunair”, “lacune”, “microvessel”, “microangiopathic”, and “small vessel”. In a new binary variable, “likely lacunar stroke”, patients were indicated as “yes” if they had mention of at least one of these keywords in the same clinic visit the year of their stroke/TIA and/or years after. Further, patients with clearly articulated non-lacunar stroke types were excluded from this count.

### Prescribing practice changes

Prescription trends of ASA or other antiplatelet use (hereafter “ASA/AP”) were analyzed among FD patients without prior ischemic MACE at representative timepoints before and after 2018 (when the ASCEND, ARRIVE and ASPREE trials were published). Patients were identified who had ASA/AP data captured in the last 3 years (2021-2023, “post-trials”) and/or in the 3 years prior to the 3 trials’ publication (2015-2017, “pre-trials”). Exclusion criteria for these analyses were: no medication records available of ASA and/or APs during any of these 6 years; any MACE ever during the study period (as treatment with ASA/AP following this event would constitute secondary prevention); or were on oral anticoagulants (e.g., warfarin, apixaban, etc.) at any point during these 6 years. Data from the most recent available year in these “pre” and “post” windows were used (e.g., if data was present in both 2022 and 2023, 2023 data was used).

### ASCVD risk

10-year ASCVD risk was calculated using the 2013 ACC/AHA guidelines^15^ via the “ascvd_10y_accaha” function in the R package “CVrisk.” Variables required for the risk calculation were: age, sex, race, total cholesterol, HDL cholesterol, systolic blood pressure, on any antihypertensive medications, smoking status, and having a diagnosis of diabetes.

For the stroke/TIA group, variables for input into the calculator were identified from the same clinic visit (year) that the patient had their first stroke/TIA, thus representing their “primary” risk at the time of their event. If a measurement wasn’t available for that year, the entry from the previous visit was used. For patients in the “primary prevention” group with data at both pre-and post-trial timepoints, an average score was calculated between the two dates, otherwise the time point (pre or post) with complete data was used.

### Statistical testing

For categorical variable significance testing, the chi-squared test (unpaired data) or McNemar’s chi-squared test (paired data) were used. Wilcoxon rank sum test was used to assess significance between groups for the numeric variables as all numeric data were non-normally distributed. Statistical significance was defined as a p value < 0.05.

## Results

### Cohort characteristics

A total of 641 patients were recruited into CFDI from 2007 through to the date of data extraction, June 2023. Out of the 641 patients, 414 (65%) were female and 227 (35%) male, in keeping with the disease’s X-linked dominant mode of inheritance. There were 203 (32%) with attenuated *GLA* variants, 407 (63%) with severe *GLA* variants, and 31 (5%) with a variant of unknown or highly variable severity. The most common variants represented in this cohort were the Nova Scotian founder variant p.Ala143Pro (n=124, 19.3%), followed by the late-onset p.Asn215Ser variant (n=57, 8.9%) and the Taiwanese founder intronic variant c.640-801G>A (n=22, 3.4%). A full list of *GLA* variants in the CFDI and severity classification are summarized in Supplemental Table S1.

Of the 641 total patients in the cohort, 120 patients (18.7%) had an ever-history of stroke/TIA, MI or PCI/CABG. Among these, the majority type event was prior stroke/TIA (n=83, 69%) followed by MI (n=28, 23%) then PCI/CABG (n=24, 20%).

There were a total of 57 patients for whom their incident stroke/TIA occurred during the study period (27 females, 30 males). Among these 57, 25 (44%) had ≥2 stroke/TIAs, for a total of 98 cumulative stroke/TIAs. Further details are described in Supplemental Table S2.

In comparing prescription trends of ASA/AP pre-2018 versus post-2018 timepoints of patients without prior MACE (“Primary Prevention Group”), 399 patients remained after exclusion criteria. Of these 399, 193 had ASA/AP use data at both timepoints, and 206 had data at only one timepoint (79 pre-2018 only and 127 post-2018 only). In this group, 141 were female (73%) and 52 (27%) male, and 137 (71%) had severe *GLA* variants, 50 (26%) had attenuated variants, and 6 (3%) had variants of unknown category.

Demographic, FD-specific characteristics, and ASCVD risk factors are summarized in Table 1 between the stroke/TIA group (n=57) and the primary prevention group with ASA/AP prescription data at both timepoints (n=193). Compared to the stroke/TIA group, in the primary prevention group there were significantly more females (p<0.001), younger age (p<0.001), lower Lyso-Gb3 (p<0.001), lower LVMI (p<0.001), higher eGFR (p<0.001), less dyslipidemia (p=0.001) less hypertension (p<0.001), and lower 10-year ASCVD risk score (p<0.001). Characteristics for the patients with only one timepoint of ASA/AP prescription data are summarized in Supplemental Table S3, which represent patients with prescription data only at “pre” or “post” timepoints.

**Table 1:**
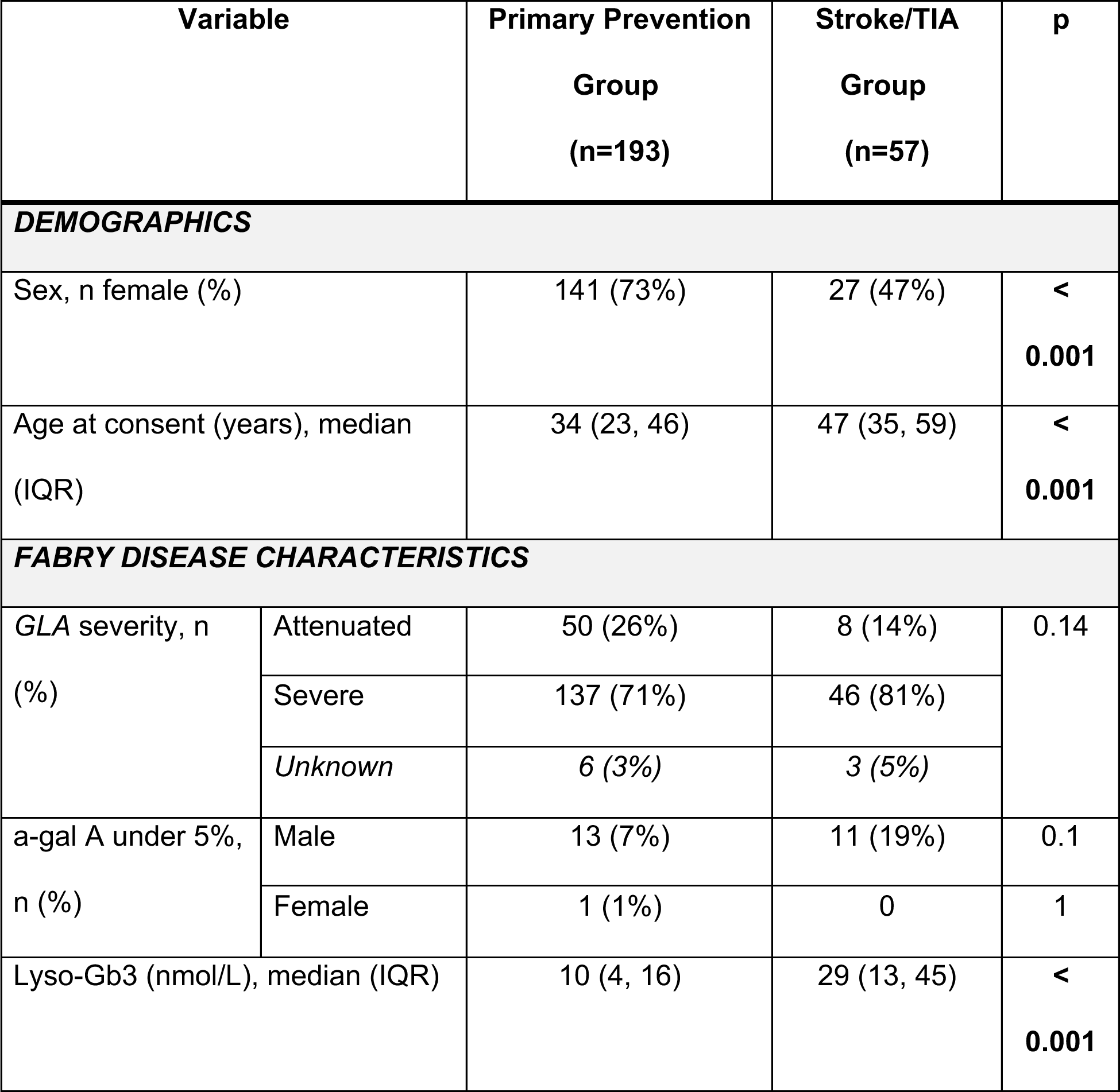
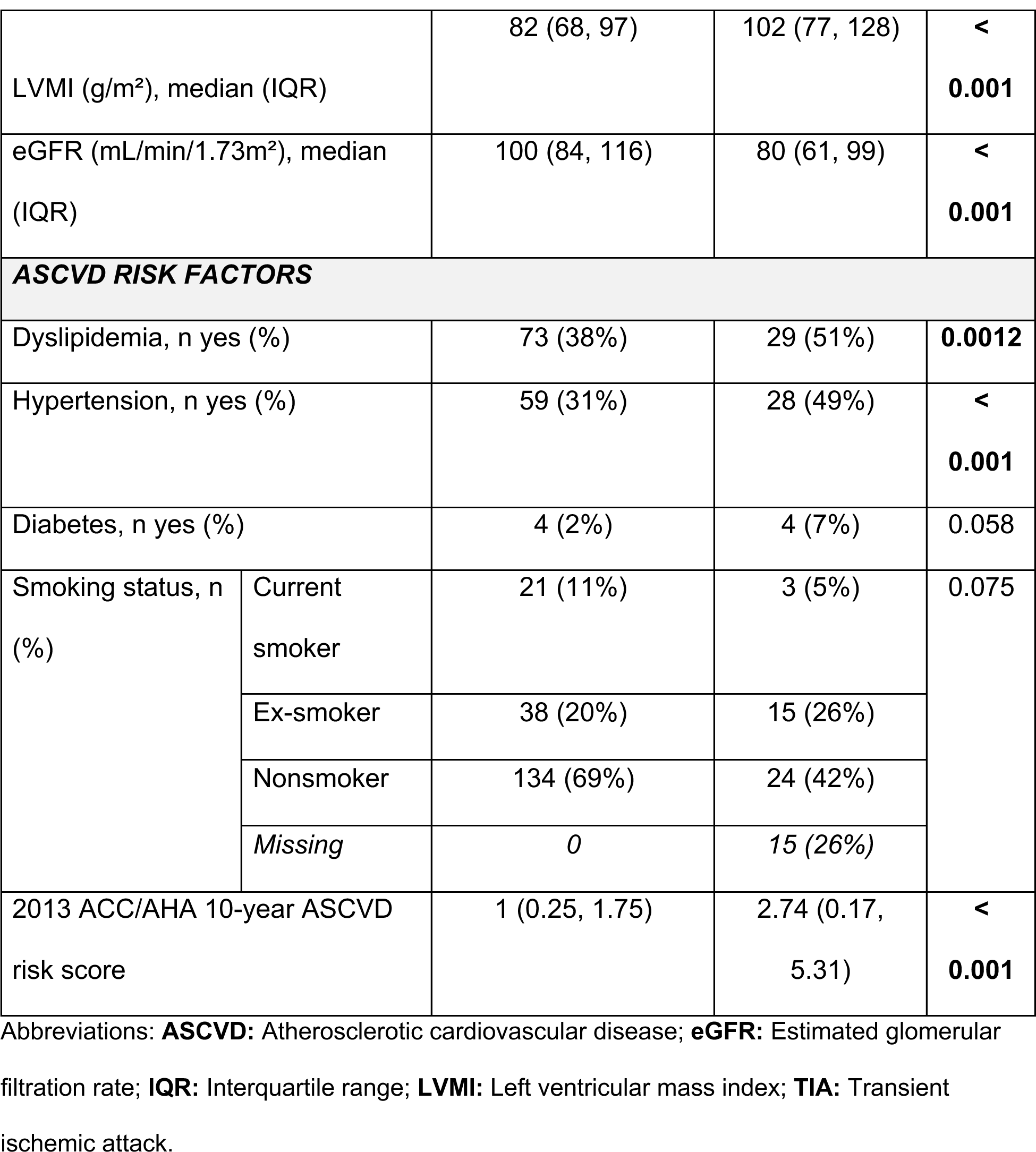
Cohort characteristics comparing patients without prior major adverse cardiovascular events and prescription data at pre-and post-trial timepoints (n=193) to patients with incident stroke/TIA during the study period (n=57).

| Variable |  | Primary Prevention<br>Group<br>(n=193) | Stroke/TIA<br>Group<br>(n=57) | p |
| --- | --- | --- | --- | --- |
| **DEMOGRAPHICS** |  |  |  |  |
| Sex, n female (%) |  | 141 (73%) | 27 (47%) | <<br>**0.001** |
| Age at consent (years), median<br>(IQR) |  | 34 (23, 46) | 47 (35, 59) | <<br>**0.001** |
| **FABRY DISEASE CHARACTERISTICS** |  |  |  |  |
| GLA severity, n<br>(%) | Attenuated | 50 (26%) | 8 (14%) | 0.14 |
|  | Severe | 137 (71%) | 46 (81%) |  |
|  | Unknown | 6 (3%) | 3 (5%) |  |
| a-gal A under 5%,<br>n (%) | Male | 13 (7%) | 11 (19%) | 0.1 |
|  | Female | 1 (1%) | 0 | 1 |
| Lyso-Gb3 (nmol/L), median (IQR) |  | 10 (4, 16) | 29 (13, 45) | <<br>**0.001** |
| LVMI (g/m<sup>2</sup>), median (IQR) |  | 82 (68, 97) | 102 (77, 128) | <<br>**0.001** |
| eGFR (mL/min/1.73m<sup>2</sup>), median (IQR) |  | 100 (84, 116) | 80 (61, 99) | <<br>**0.001** |
| **ASCVD RISK FACTORS** |  |  |  |  |
| Dyslipidemia, n yes (%) |  | 73 (38%) | 29 (51%) | **0.0012** |
| Hypertension, n yes (%) |  | 59 (31%) | 28 (49%) | <<br>**0.001** |
| Diabetes, n yes (%) |  | 4 (2%) | 4 (7%) | 0.058 |
| Smoking status, n (%) | Current smoker | 21 (11%) | 3 (5%) | 0.075 |
|  | Ex-smoker | 38 (20%) | 15 (26%) |  |
|  | Nonsmoker | 134 (69%) | 24 (42%) |  |
|  | *Missing* | *0* | *15 (26%)* |  |
| 2013 ACC/AHA 10-year ASCVD risk score |  | 1 (0.25, 1.75) | 2.74 (0.17, 5.31) | <<br>**0.001** |

Abbreviations: **ASCVD**: Atherosclerotic cardiovascular disease; **eGFR**: Estimated glomerular filtration rate; **IQR**: Interquartile range; **LVMI**: Left ventricular mass index; **TIA**: Transient ischemic attack.

### Stroke/TIA incidence and characteristics

The overall stroke/TIA rate per patient year was 0.016. Males with severe variants had an overall stroke/TIA incidence rate approximately three times that of females of either variant severity type or males with attenuated variant. Males had an overall stroke/TIA rate 2.65 times that of females (0.026 vs. 0.0098). Males with severe variants drove most of this variation (0.031), whereas males with attenuated variants had overall stroke rates (0.011) similar to females with severe variants (0.0096). Overall, males had an incident stroke rate 2.1 times that of females (0.014 vs 0.0066). Interestingly, the incident stroke rate in males with attenuated variants (0.008) was similar to that of females with severe variants (0.0076). These results are further detailed in Supplemental Table S4.

Incident stroke/TIA rate by sex and *GLA* showed marked differences between age groups (<40, 50-59 and 60+) (Figure 1). During the study period, 34 (60%) of the 57 patients were under 60 years old at the time of their incident stroke. There were 24 patients who had their first stroke/TIA between ages 40-59, the age range within which recommendations for ASA/AP for primary prevention are debated. Out of these 24, 21 (88%) had severe variants, 2 (8%) were attenuated and 1 (4%) unknown. Incident stroke/TIA rates by sex, *GLA* and age group are summarized in Supplemental Table S5.

**Figure 1:**
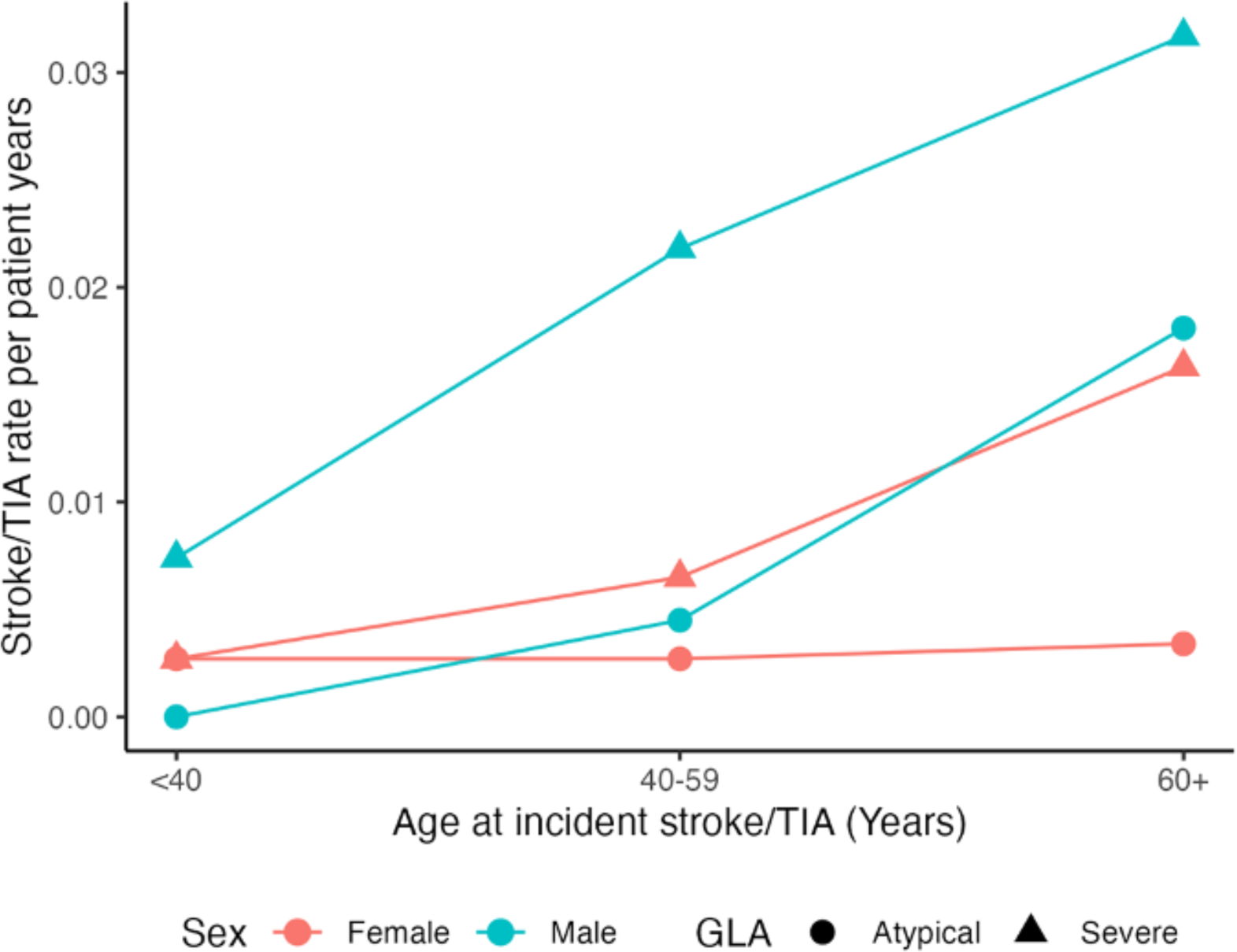
Incident stroke/transient ischemic attack (TIA) rate per patient years by sex and *GLA* variant severity and age group of incident event (<40, 40-59 or 60+ years of age).

There were notable differences between the incident stroke/TIA rate observed in the CFDI FD cohort compared to the general Canadian statistics: FD females had consistently around 9 times higher incident rates than non-FD Canadian women, and FD males had dramatically higher rates compared to non-FD Canadian men (Figure 2). Stroke/TIA rates are summarized in Supplemental Table S6.

**Figure 2:**
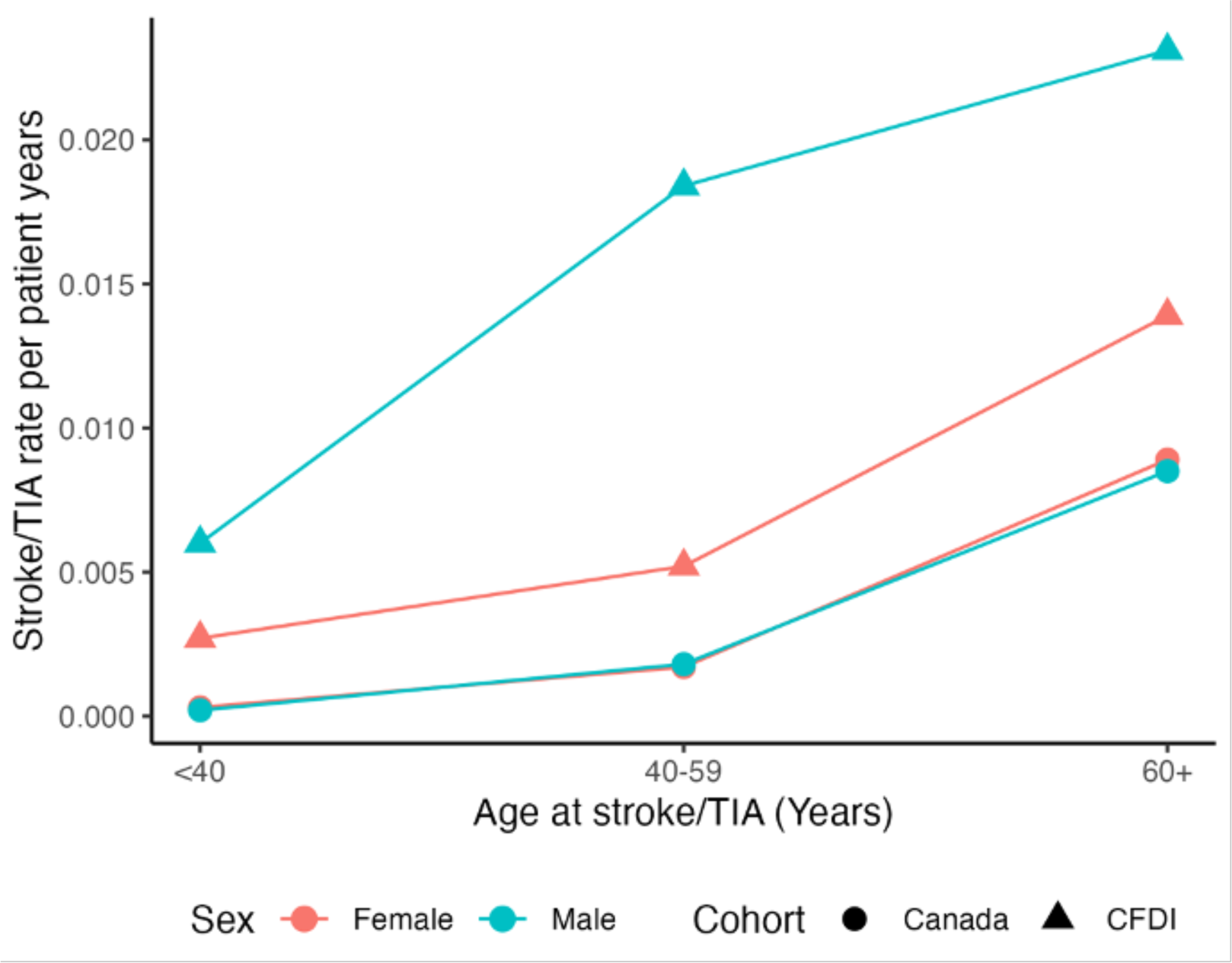
Incident stroke/transient ischemic attack (TIA)* rate per patient by sex and cohort (Canadian Fabry Disease Initiative (CFDI) versus 2012/13 Canadian stroke statistics). **\***Canadian statistics were stroke only (not TIA) and did not distinguish incident from recurrent strokes.

Of the 57 patients under age 60 years at the time of incident stroke, 41 patients (72%) were on enzyme replacement therapy (ERT) or molecular chaperone therapy (Migalastat) prior to their incident stroke/TIA event. The majority were on ERT (39/41, 95%) compared to chaperone (2/41, 5%). Of those on ERT, 30 were on agalsidase alfa immediately prior to their stroke/TIA, and 9 were on agalsidase beta.

Review of qualitative stroke/TIA descriptions resulted in 30/57 patients (53%) with suggestions of lacunar stroke. Of these 57 patients, 16 (28%) were on an anticoagulant in the year prior to their incident stroke/TIA. A limitation is that, for most patients, it wasn’t possible with available data to distinguish stroke/TIA subtypes even if small vessel disease was observed (i.e., if large-vessel atherosclerotic/embolic etiology was excluded).

### ASA/AP prescribing trends

Among the patients in the primary prevention group ever on antiplatelet therapy without anticoagulants, most patients were on ASA monotherapy (185/193, 96%); the rest (n=8, 4%) were on both ASA and another AP, or just another AP (dipyridamole, clopidogrel, ticagrelor or ticlidopine). When comparing ASA/AP prescribing trends between pre-and post-2018 timepoints, there was a significant (p=0.01) absolute decrease of 8% in the percentage of patients on ASA/AP for primary prevention from 86 (45%) to 72 (37%) (Table 2).

**Table 2:**
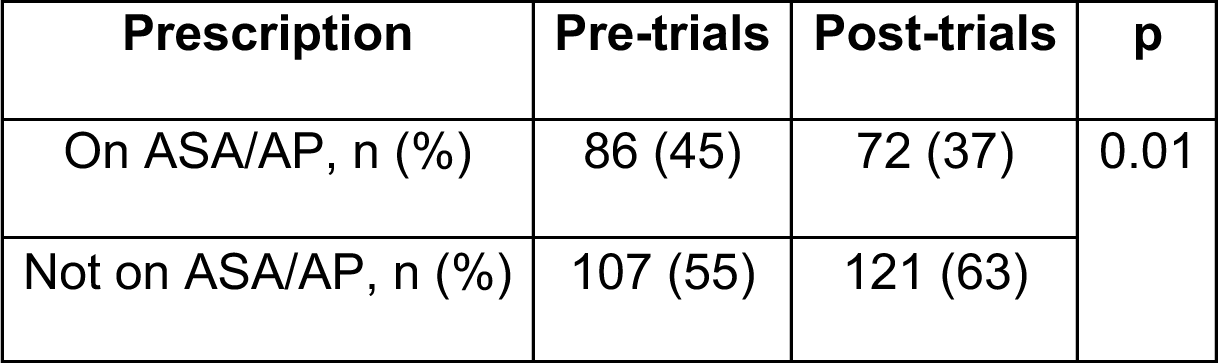
Prescription trends in the patients on acetylsalicylic acid (ASA) and/or antiplatelet (AP) therapy (n=193) between pre-trial and post-trial timepoints.

| **Prescription** | **Pre-trials** | **Post-trials** | **p** |
| --- | --- | --- | --- |
| On ASA/AP, n (%) | 86 (45) | 72 (37) | 0.01 |
| Not on ASA/AP, n (%) | 107 (55) | 121 (63) |  |

Prescription trends between the two timepoints were as follows: 54 (28%) stayed on ASA/AP, 32 (17%) were de-prescribed ASA/AP, 18 (9%) were prescribed ASA/AP, and 89 (46%) were never on ASA/AP. Prescription trends were compared according to *GLA* variant severity (severe or attenuated, n=187) and by sex (male or female, n=193). Significant differences were observed by *GLA* variant (p=0.013) but not by sex (p=0.60), most notably in that more patients with attenuated *GLA* variants were deprescribed ASA/AP (32%) than patients with severe variants (12%). These results are summarized in Supplemental Table S7.

### Cardiovascular risk between groups

A majority (n=165, 85%) of the 193 patients in the primary prevention group had the required data needed to calculate at least one 10-year ASCVD risk score using the 2013 ACC/AHA risk calculator. None of the patients under 60 years old at the time of their first stroke/TIA would have been considered moderate-to-high (≥10%) 10-year ASCVD risk based on this calculator alone (Figure 3). Of the 23 patients 60 years or older, 12/23 (52%) had moderate-to-high ASCVD risk. Notably, none of these older 23 patients ever had recorded PCI/CABG, and only one had MI (with no PCI/CABG) prior to their first event.

**Figure 3:**
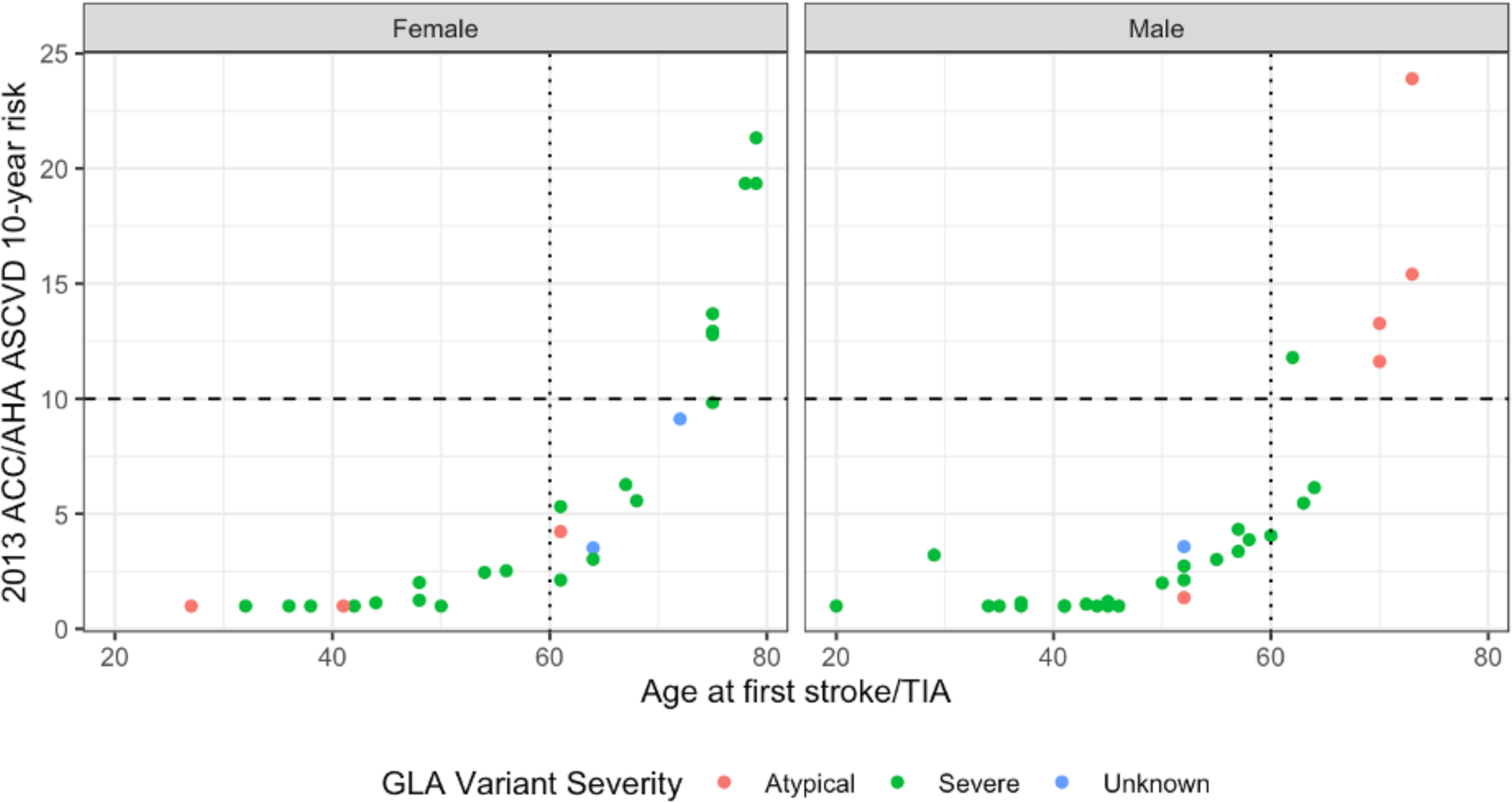
10-year 10% risk score of atherosclerotic cardiovascular disease (ASCVD) among patients with prior stroke/transient ischemic attack (TIA) (n=57). Risk score of patients across age groups using the 2013 ACC/AHA ASCVD 10-year risk calculator using variables recorded at the year of their first stroke/TIA.

History of preceding PCI with stent or CABG was investigated for all 57 patients with incident stroke/TIA during the study period as a correlate to suggest potentially atherosclerotic origin for their event. 5/57 (8.8%) ever had a history of PCI/CABG. Only 1/5 had preceding MI to the PCI/CABG. 2/5 patients had their first stroke/TIAs preceding their PCI/CABG, 2/5 had PCI/CABG preceding their first stroke/TIAs, and 1/5 had their PCI/CABG the same year as their first stroke/TIA. 3/5 of these patients had qualitative notes in brain MRI/CT imaging that suggested lacunar stroke/TIA. Further details are summarized in Supplemental Table S8.

## Discussion

Our cohort of FD patients displayed a high incidence of stroke/TIA relative to the Canadian population, especially at younger ages when their 10-year ASCVD risk predictions were low and unhelpful (<<10%). A higher rate of stroke/TIAs was observed among FD males than females, and predominantly patients with severe *GLA* variants for both sexes. Risk management strategies for stroke/TIA risk in FD patients should take both sex and *GLA* variant severity into consideration, should any effective prevention strategies be identified in the future.

The 2013 ACC/AHA ASCVD risk calculators weigh older age and presence of diabetes more heavily than other variables, which may explain the relatively low ASCVD risk percentages observed despite higher prevalence of dyslipidemia, hypertension and smoking in the stroke/TIA group compared to the primary prevention group. The 2022 USPSTF advisory statement^13^ advises personalized discretion for prescription of ASA for primary prevention of MACEs in the 40-59-year-old age group of the general population if they have ≥10% ASCVD risk as calculated by the 2013 ACC/AHA guidelines^15^. This advisory statement advises that, in the general population, evidence for net benefit of ASA use in this age group is limited even if in the group with ≥10% 10-year risk. In comparison, the presence of FD alone (especially in a male) as a risk factor overwhelms the utility of ASCVD risk calculators in risk stratifying patients for stroke prevention.

A majority of FD patients had suggestions of lacunar etiology (30/57, 53%), however, more precise documentation of the subtype of stroke/TIA events in the registry was lacking, and the true portion with small vessel strokes is likely higher. There was some documentation on the presence or absence of white matter lesions in the brain, however after considering concerns about the robustness of the information available, these data were excluded. The observations that most FD patients with stroke/TIA never had evidence of obstructive coronary artery disease while in the study (52/57, 91%), and that most had <10% 10-year ASCVD risk at the time of their first stroke/TIA (46/57, 81%) suggests atherosclerotic sources causing thrombotic cerebrovascular clotting events may not be the predominant pathophysiology of stroke/TIA in these early-presenting FD patients.

Two large studies of FD patients describe lacunar and cryptogenic stroke as the most common etiologies in FD^5,7^. However, there is currently no standardized prophylactic therapy recommended for cerebral small vessel disease. In the Fabry Registry of 2,446 patients, lacunar (small vessel) stroke was the most common ischemic etiology among their patients (70.4%)^5^. Lacunar stroke is a marker of cerebral small vessel disease, and is defined as a subcortical infarct measuring less than 20 mm in diameter, which is the end result of liquefactive necrosis caused by occlusion of a perforator vessel from an intracranial artery. Mechanisms that can cause lacunar stroke include atherosclerotic disease, cardiac embolism, and lipohyalinosis, among which lipohyalinosis is the most common lacunar stroke mechanism^16^. Lipohyalinosis is a concentric hyaline thickening of the cerebral small vessels leading to the occlusion of the small penetrating arteries. Further, endothelial dysfunction and impaired autoregulation in the microvasculature have been demonstrated to be important contributing factors to the development of lacunar infarcts^17^. Second, Ortiz et al.^7^ meta-analyzed the prevalence of cryptogenic or first-time strokes in 1,986 FD patients across 11 cohort studies. The consensus was that the increased likelihood of cryptogenic or first-stroke in these FD patients was primarily due to disturbances in vasoreactivity and autoregulation in the vascular endothelium as a result of the accumulation of glycosphingolipids in those cells, suggesting vasospasm^18^, reduced cerebral blood flow velocities and impaired autoregulation^19^, and upregulation of angiotensin II^20^ as key contributing mechanisms. Recommendations of antithrombotic therapy for primary and secondary prevention of ischemic stroke vary depending on the etiology of the event. ASA has been indicated as the recommended single antiplatelet therapy for large-artery atherosclerosis or embolic cryptogenic strokes, and oral anticoagulants are recommended for cardioembolic strokes^21^. An uncertain benefit of cilostazol has been suggested for lacunar stroke, which is an agent that exhibits a primarily vasodilatory function and some antiplatelet activity^21^. Hou et al.^22^ discuss the efficacy of cilastazol over other antiplatelets such as ASA for secondary prevention of any cerebrovascular or cardiovascular event after lacunar stroke, stating that there is insufficient evidence that these other antiplatelets can directly affect the presumed lipohyalinotic mechanism of most lacunar strokes. In contrast, Kwok et al.^23^ support the use of ASA for secondary stroke prevention after a lacunar stroke, with the presumption of lacunar stroke etiology arising from arteriosclerosis in the deeper arteries.

An interpretation of the conflicting suggestions about ASA’s efficacy in preventing incident stroke/TIA events, in the context of FD patients, pertains to the inability to precisely subcategorize lacunar and cryptogenic strokes, the most common etiologies in FD, as atherosclerotic/embolic in origin (ASA more beneficial) than lipohyalinotic, non-atherosclerotic or non-embolic in etiology (ASA possibly less beneficial).

There is conflicting evidence of any benefit from ERT or chaperone towards prevention of incident stroke/TIA. If any benefit exists, it is insufficient, as most of the patients were on therapy at the time of their incident event. A focus of future FD therapy development should include sustained and robust drug entry into vascular endothelial cells. In addition, we propose that cerebrovascular endpoints, such as white matter lesion burden on imaging, should be explored as a possible outcome measure in clinical trials.

### Limitations

It is unlikely that a well-powered randomized controlled trial will ever be performed in FD to test for the benefit of ASA/AP. Even our large registry was not of sufficient size to retrospectively assess for associations of risk-lowering while also controlling for age, disease severity, and comorbidities. Another limitation of the registry was the lack of data capturing major bleeding events in the population, which are the major adverse reactions associated with ASA. Lastly, atrial fibrillation is a known risk factor for embolic stroke, however this diagnosis was not systematically collected. Although 28% of patients were anticoagulated prior to their incident stroke/TIA, we cannot conclude definitively that these patients had atrial fibrillation, as they may have been anticoagulated for other reasons; further, some patients may have atrial fibrillation and not have been anticoagulated due to a prior contraindication (e.g., prior bleeding events).

## Data Availability

Summary data produced in the present study are available upon reasonable request to the authors.

## Non-standard Abbreviations and Acronyms

a-gal A: Alpha-galactosidase A
AP: Antiplatelet
ASA: Acetylsalicylic acid
ASCVD: Atherosclerotic cardiovascular disease
CABG: Coronary artery bypass grafting
CFDI: Canadian Fabry Disease Initiative
CT: Computed tomography
CVD: Cardiovascular Disease
eGFR: Estimated glomerular filtration rate
ERT: Enzyme replacement therapy
FD: Fabry disease
Gb3: Globotriaosylceramide
GLA: Galactosidase alpha
HF: Heart failure
LVMI: Left ventricular mass index
Lyso-Gb3: Globotriaosylsphingosine
MACE: Major adverse cardiovascular event
MI: Myocardial Infarction
MRI: Magnetic resonance imaging
PCI: Percutaneous coronary intervention
TIA: Transient ischemic attack
USPSTF: United States Preventive Service Task Force

## Acknowledgments

The authors would like to thank the CFDI patients and their families for their participation in this registry; Dr. Karin Humphries for statistical consultation; research coordinators, nurses, including Wendy Paquin, physicians, and allied health staff in ongoing contribution to the CFDI registry.

