## Supplemental Material for "Stroke rates, risk factors, and aspirin prescribing trends in the Canadian Fabry Disease Initiative cohort"

#### CFDI Investigator Group Details

All investigators below (MD, FRCPC) oversee the active sites in Canada who are conducting ongoing recruitment and follow-up for patients followed in the CFDI.

Halifax, Nova Scotia: Dr. Michael West, Dr. Sarah Dyack
Corner Brook, Newfoundland: Dr. Stephen Murphy
St. John's, Newfoundland: Dr. Marissa Chard, Dr. Lesley Turner
Moncton, New Brunswick: Dr. Alier Marrero
Montreal, Quebec: Dr. Daniel Bichet
Sherbrooke, Quebec: Dr. Bruno Maranda
Quebec City, Quebec: Dr. Paul Isenring
Toronto, Ontario: Dr. Mark Iwanochko, Dr. Chantal Morel, Dr. Michal Inbar-Feigenberg
Kingston, Ontario: Dr. Jennifer MacKenzie, Dr. Jocelyn Garland
London, Ontario: Dr. Natalya Karp, Dr. Chitra Prasad
Hamilton, Ontario: Dr. Matthew Lanktree
Ottawa, Ontario: Dr. Andrea Yu
Calgary, Alberta & Saskatoon, Saskatchewan: Dr. Aneal Khan
Edmonton, Alberta: Dr. Alicia Chan, Dr. Shailly Jain
Winnipeg, Manitoba: Dr. Cheryl Greenberg, Dr. Aziz Mhanni
Vancouver, British Columbia: Dr. Anna Lehman

#### Supplemental Methods

The pathogenic *GLA* variant was categorized for each patient as "severe" or "attenuated" for these analyses. Other groups have used a-gal A enzyme cut-offs of 1%, 1.5%, 2%, 5%, or even 30% as a cut-off for defining a "classical disease" prediction (eg., Nowak et al.[24], Schiffman et al.[25]); to avoid further inconsistency in defining "classical disease/variants," we elected to use the terms severe and attenuated. Variant descriptions were standardized according to the most recent human genome variant HGVS rules, and variant sub-grouping was determined using a sequential rationale of 4 criteria below:

1. If the variant was predicted to result in total loss of function (eg., frameshifting or truncating variant), then we automatically assigned it as a severe variant. If it was not predicted total loss of function, the second criteria was employed.
2. If the variant was represented in the Fabry GenPhen database (fabrygenphen.com), variant categorization was determined based on the annotation for males. If the variant was listed in the database as "classical", the variant was categorized as severe; if listed as "nonclassical", the variant was categorized as attenuated; if listed as "benign", subsequent criteria was consulted. Furthermore, if males had both classical and nonclassical entries

listed, then the category represented by the majority was used. If the variant only had female entries, then the third criteria was employed.

3. If the variant was not in GenPhen and in a male participant, we used an arbitrary cut-off of enzyme activity of <5% of the mean of the normal range to assign severity status from their own testing. If the participant was female, we used the average of enzyme activities from other male participants with that variant, again with a 5% activity cut-off. If the participant was female and there were no male participants with that variant, the fourth criteria was employed.
4. The literature was further searched for a report on enzyme activity in a male, using sources such as the Fabry Mutant Database (fabry-database.org) and cited papers followed up on via PubMed to ensure correct interpretation and annotation of the variant.

Failing the above rules, we listed the variant's severity status as unknown.

### Supplemental Tables

**Supplemental Table S1:** GLA variants in CFDI and ascribed severity classification.

| **Unique (117)** | **Count** | **% of 641** | **Classification (Severe, attenuated or unknown)** | **Type of variant (missense, intronic, deletion, duplication, insertion)** | **Criteria used (1-4) (1: Total Loss of Function (LoF); 2: Genphen; 3: Male enzyme cutoffs; 4: Literature)** |
|---|---|---|---|---|---|
| c.427G>C (p.Ala143Pro) | 124 | 19.3 % | Severe | Missense | 2.Genphen |
| c.644A>G (p.Asn215Ser) | 57 | 8.9% | Attenuated | Missense | 2. Genphen |
| c.640-801G>A | 22 | 3.4% | Attenuated | Intronic | 2. Genphen |
| c.1066C>T (p.Arg356Trp) | 21 | 3.3% | Attenuated | Missense | 2. Genphen |
| c.1042G>C (p.Ala348Pro) | 20 | 3.1% | Severe | Missense | 2. Genphen |
| c.1046G>A (p.Trp349*) | 16 | 2.5% | Severe | Missense | 1. Total LoF |

| | | | | | |
|---|---|---|---|---|---|
| c.1033T>C (p.Ser345Pro) | 14 | 2.2% | Severe | Missense | 2. Genphen |
| c.370-533_c.1277del4.5kb | 13 | 2.0% | Severe | Deletion | 1. Total LoF |
| c.658C>T (p.Arg220*) | 13 | 2.0% | Severe | Missense | 1. Total LoF |
| c.572T>A (p.Leu191Gln) | 13 | 2.0% | Severe | Missense | 2. Genphen |
| c.334C>T (p.Arg112Cys) | 12 | 1.9% | Severe | Missense | 2. Genphen |
| c.1024C>T (p.Arg342*) | 12 | 1.9% | Severe | Missense | 1. Total LoF |
| c.401A>C (p.Tyr134Ser) | 11 | 1.7% | Attenuated | Missense | 2. Genphen |
| c.961C>G (p.Gln321Glu) | 11 | 1.7% | Severe | Missense | 2. Genphen |
| c.854C>A (p.Ala285Asp) | 11 | 1.7% | Attenuated | Missense | 2. Genphen |
| Unknown | 11 | 1.7% | Unknown | Unknown | - |
| c.335G>A (p.Arg112His) | 10 | 1.6% | Attenuated | Missense | 2. Genphen |
| c.1156C>T (p.Gln386*) | 9 | 1.4% | Severe | Missense | 1. Total LoF |
| c.352C>T (p.Arg118Cys) | 9 | 1.4% | Attenuated | Missense | 3. Male enzyme cutoffs |
| c.680G>A (p.Arg227Gln) | 8 | 1.2% | Severe | Missense | 2. Genphen |
| c.1025G>A (p.Arg342Gln) | 7 | 1.1% | Severe | Missense | 2. Genphen |
| c.877C>A (p.Pro293Thr) | 7 | 1.1% | Attenuated | Missense | 3. Male enzyme cutoffs |
| c.496_497delins (p.Leu166Ser) | 7 | 1.1% | Unknown | Deletion + Insertion (no frameshift) | - |
| c.369+5G>T | 6 | 0.9% | Attenuated | Intronic | 2. Genphen |
| c.1088G>A (p.Arg363His) | 6 | 0.9% | Attenuated | Missense | 2. Genphen |
| c.994del (p.Arg332Aspfs*16) | 6 | 0.9% | Severe | Deletion | 1. Total LoF |
| c.962A>G (p.Gln321Arg) | 5 | 0.8% | Attenuated | Missense | 2. Genphen |

| | | | | | |
|---|---|---|---|---|---|
| c.802-3_804delins | 5 | 0.8% | Severe | Deletion | 3. Male enzyme cutoffs |
| c.1241T>C (p.Leu414Ser) | 5 | 0.8% | Severe | Missense | 2. Genphen |
| c.761_763del (p.Val254del) | 5 | 0.8% | Severe | Deletion | 4. Literature |
| c.613C>A (p.Pro205Thr) | 5 | 0.8% | Attenuated | Missense | 2. Genphen |
| c.277del (p.Asp93Thrfs*28) | 5 | 0.8% | Severe | Deletion | 1. Total LoF |
| c.386T>C (p.Leu129Pro) | 4 | 0.6% | Severe | Missense | 2. Genphen |
| c.427G>A (p.Ala143Thr) | 4 | 0.6% | Attenuated | Missense | 4. Literature |
| c.1201dup (p.Ser401Phefs*50) | 4 | 0.6% | Severe | Duplication | 1. Total LoF |
| c.137A>C (p.His46Pro) | 4 | 0.6% | Severe | Missense | 3. Male enzyme cutoffs |
| c.1012G>A (p.Glu338Lys) | 4 | 0.6% | Attenuated | Missense | 2. Genphen |
| c.679C>T (p.Arg227*) | 4 | 0.6% | Severe | Missense | 1. Total LoF |
| c.707G>T (p.Trp236Leu) | 3 | 0.5% | Severe | Missense | 2. Genphen |
| c.317_327del (p.Leu106Profs*13) | 3 | 0.5% | Severe | Deletion | 1. Total LoF |
| c.744_745del (p.Phe248Leufs*7) | 3 | 0.5% | Severe | Deletion | 1. Total LoF |
| c.266T>C (p.Leu89Pro) | 3 | 0.5% | Attenuated | Missense | 2. Genphen |
| c.1000-2A>G | 3 | 0.5% | Severe | Intronic | 2. Genphen |
| c.956T>C (p.Ile319Thr) | 3 | 0.5% | Severe | Missense | 2. Genphen |
| c.1115T>G (p.Leu372Arg) | 3 | 0.5% | Attenuated | Missense | 2. Genphen |
| c.1139C>T (p.Pro380Leu) | 3 | 0.5% | Attenuated | Missense | 3. Male enzyme cutoffs |
| c.105dup (p.Leu36Ilefs*20) | 3 | 0.5% | Severe | Duplication | 1. Total LoF |
| c.902G>A (p.Arg301Gln) | 3 | 0.5% | Attenuated | Missense | 4. Literature |

| | | | | | |
|---|---|---|---|---|---|
| c.1072G>A (p.Glu358Lys) | 3 | 0.5% | Severe | Missense | 4. Literature |
| c.242G>A (p.Trp81*) | 3 | 0.5% | Severe | Missense | 1. Total LoF |
| c.128G>T (p.Gly43Val) | 3 | 0.5% | Attenuated | Missense | 2. Genphen |
| c.612G>A (p.Trp204*) | 3 | 0.5% | Severe | Missense | 1. Total LoF |
| c.878C>T (p.Pro293Leu) | 2 | 0.3% | Severe | Missense | 2. Genphen |
| c.2T>C (p.Met1Thr) | 2 | 0.3% | Severe | Missense | 2. Genphen |
| c.869T>C (p.Met290Thr) | 2 | 0.3% | Attenuated | Missense | 3. Male enzyme cutoffs |
| c.833dup (p.Asn278Lysfs*21) | 2 | 0.3% | Severe | Duplication | 1. Total LoF |
| c.295del (p.Gln99Lysfs*22) | 2 | 0.3% | Severe | Deletion | 1. Total LoF |
| c.782G>T (p.Gly261Val) | 2 | 0.3% | Severe | Missense | 4. Literature |
| c.774_775delAC (p.Pro259Argfs*5) | 2 | 0.3% | Severe | Deletion | 1. Total LoF |
| c.506T>C (p.Phe169Ser) | 2 | 0.3% | Severe | Missense | 4. Literature |
| c.729G>C (p.Leu243Phe) | 2 | 0.3% | Severe | Missense | 3. Male enzyme cutoffs |
| c.299G>C (p.Arg100Thr) | 2 | 0.3% | Severe | Missense | 4. Literature |
| c.374A>C (p.His125Pro) | 2 | 0.3% | Unknown | Missense | - |
| c.621dup (p.Met208Tyrfs*24) | 2 | 0.3% | Severe | Duplication | 1. Total LoF |
| c.692A>C (p.Asn231Gly) | 2 | 0.3% | Severe | Missense | 2. Genphen |
| c.195-?547?+?del (Exons 2 and 3 deleted) | 2 | 0.3% | Severe | Deletion | 1. Total LoF |
| c.961C>T (p.Gln321*) | 2 | 0.3% | Severe | Missense | 1. Total LoF |
| c.233C>G (p.Ser78*) | 2 | 0.3% | Severe | Missense | 1. Total LoF |
| c.962A>T (p.Gln321Leu) | 2 | 0.3% | Attenuated | Missense | 2. Genphen |
| c.394G>A (p.Gly132Arg) | 2 | 0.3% | Unknown | Missense | - |

| | | | | | |
|---|---|---|---|---|---|
| c.1288del (p.Ter430Lysext*?) | 2 | 0.3% | Severe | Deletion | 1. Total LoF |
| c.901C>G (p.Arg301Gly) | 2 | 0.3% | Severe | Missense | 4. Literature |
| c.1082G>T (p.Gly361Val) | 2 | 0.3% | Severe | Missense | 3. Male enzyme cutoffs |
| c.559_560del (p.Met187Valfs*6) | 2 | 0.3% | Severe | Deletion | 1. Total LoF |
| c.618_621delins (p.Tyr207Lysfs*33) | 2 | 0.3% | Severe | Deletion + Insertion (frameshift) | 1. Total LoF |
| c.893_894ins (p.Asn298Lysfs*2) | 2 | 0.3% | Severe | Insertion | 1. Total LoF |
| c.242G>C (p.Trp81Ser) | 2 | 0.3% | Severe | Missense | 2. Genphen |
| c.234del (p.Glu79Lysfs*42) | 1 | 0.2% | Severe | Deletion | 1. Total LoF |
| c.774del (p.Pro259Glnfs*10) | 1 | 0.2% | Severe | Deletion | 1. Total LoF |
| c.270dup (p.Ile91Hisfs*2) | 1 | 0.2% | Severe | Duplication | 1. Total LoF |
| c.640-2A>C (p.Exon5del) | 1 | 0.2% | Severe | Deletion | 1. Total LoF |
| c.695T>C (p.Ile232Thr) | 1 | 0.2% | Attenuated | Missense | 2. Genphen |
| c.101dup (p.Asn34Lysfs*22) | 1 | 0.2% | Severe | Duplication | 1. Total LoF |
| c.189C>A (p.Cys63*) | 1 | 0.2% | Severe | Missense | 1. Total LoF |
| c.139T>G (p.Trp47Gly) | 1 | 0.2% | Severe | Missense | 4. Literature |
| c.466G>A (p.Ala156Thr) | 1 | 0.2% | Unknown | Missense | - |
| c.1087C>T (p.Arg363Cys) | 1 | 0.2% | Attenuated | Missense | 2. Genphen |
| c.937G>T (p.Asp313Tyr) | 1 | 0.2% | Unknown | Missense | - |
| c.1181?>C (p.Leu394Pro) | 1 | 0.2% | Attenuated | Missense | 3. Male enzyme cutoffs |
| c.823C>T (p.Leu275Phe) | 1 | 0.2% | Unknown | Missense | - |
| c.776C>T (p.Pro259Leu) | 1 | 0.2% | Severe | Missense | 4. Literature |
| c.194+1562_370-892del | 1 | 0.2% | Unknown | Intronic deletion | - |
| c.422C>T (p.Thr141Ile) | 1 | 0.2% | Unknown | Missense | - |

| | | | | | |
|---|---|---|---|---|---|
| c.195-1G>A | 1 | 0.2% | Severe | Intronic | 4. Literature |
| c.272T>C (p.Ile91Thr) | 1 | 0.2% | Severe | Missense | 3. Male enzyme cutoffs |
| c.613C>T (p.Pro205Ser) | 1 | 0.2% | Attenuated | Missense | 4. Literature |
| c.797A>C (p.Asp266Ala) | 1 | 0.2% | Unknown | Missense | - |
| c.253G>A (p.Gly85Met) | 1 | 0.2% | Severe | Missense | 3. Male enzyme cutoffs |
| c.548-1G>A | 1 | 0.2% | Severe | Intronic | 4. Literature |
| c.812G>C (p.Gly271Ala) | 1 | 0.2% | Severe | Missense | 2. Genphen |
| c.1033_1034del (p.Ser345Argfs*29) | 1 | 0.2% | Severe | Deletion | 1. Total LoF |
| c.1049del (p.Ala350Valfs*2) | 1 | 0.2% | Severe | Deletion | 1. Total LoF |
| c.641C>T (p.Pro214Leu) | 1 | 0.2% | Unknown | Missense | - |
| c.775C>T (p.Pro259Ser) | 1 | 0.2% | Attenuated | Missense | 4. Literature |
| c.950T>C (p.Ile317Thr) | 1 | 0.2% | Severe | Missense | 2. Genphen |
| c.901C>T (p.Arg301*) | 1 | 0.2% | Severe | Missense | 1. Total LoF |
| c.1072_1074del (p.Glu358del) | 1 | 0.2% | Severe | Deletion | 1. Total LoF |
| c.486G>A (p.W162*) | 1 | 0.2% | Severe | Missense | 1. Total LoF |
| c.511G>C (p.Gly171Arg) | 1 | 0.2% | Unknown | Missense | - |
| c.161T>C (p.Leu54Pro) | 1 | 0.2% | Attenuated | Missense | 2. Genphen |
| c.317T>G (p.Leu106Arg) | 1 | 0.2% | Severe | Missense | 4. Literature |
| c.802-3_802-2del | 1 | 0.2% | Severe | Deletion | 4. Literature |
| c.136C>T (p.His46Tyr) | 1 | 0.2% | Unknown | Missense | - |
| c.1121_1123del (p.Lys374_Gly375delinsArg) | 1 | 0.2% | Severe | Deletion + Insertion (no frameshift) | 3. Male enzyme cutoffs |
| c.35_47del (p.Cys12Phefs*105) | 1 | 0.2% | Severe | Deletion | 1. Total LoF |

| | | | | | |
|---|---|---|---|---|---|
| c.815A>G (p.Asn272Ser) | 1 | 0.2% | Severe | Missense | 4. Literature |

**Supplemental Table S2:** Quantifying the number of stroke/TIA events during the study period (2007-2023).

| **Number of strokes/TIAs within study period** | **Total (n=641)** | **Female (n=414)** | **Male (n=227)** |
|---|---|---|---|
| 0 | 584 (91%) | 387 (93%) | 197 (87%) |
| 1 | 32 (5%) | 18 (4%) | 14 (6%) |
| 2 | 18 (3%) | 7 (2%) | 11 (5%) |
| 3 | 2 (0.3%) | 0 | 2 (0.9%) |
| 4 | 4 (0.6%) | 2 (0.5%) | 2 (0.9%) |
| 8 | 1 (0.2%) | 0 | 1 (0.4%) |

**Supplemental Table S3:** Cohort characteristics of the patients with data represented at only the pre-clinical trial timeframe (2015-2017) or just the post-clinical trial timeframe (2021-2023). Significance testing between differences of the pre- vs post-only.

| **Variable** | | **All with one timepoint only (n=206)** | **Pre-only (n=79)** | **Post-only (n=127)** | **p** |
|---|---|---|---|---|---|
| ***DEMOGRAPHICS*** | | | | | |
| Sex, n female (%) | | 141 (68%) | 50 (63%) | 91 (72%) | 0.27 |
| Age at consent (years), median (IQR) | | 31 (17, 48) | 26 (15, 37) | 39 (23, 55) | **3.91E-05** |
| ***FABRY CHARACTERISTICS*** | | | | | |
| GLA severity, n (%) | Attenuated | 80 (39%) | 26 (33%) | 54 (43%) | 0.08 |
| | Severe | 113 (55%) | 52 (66%) | 61 (48%) | |
| | *Unknown** | *13 (6%)* | *1 (1%)* | *12 (9%)* | |
| a-GAL A under 5%, n (%) | Male | 13 (6%) | 5 (6%) | 8 (6%) | 1 |
| | Female | 0 | 0 | 0 | n/a |
| Lyso-Gb3 (nmol/L), median (IQR) | | 8 (2, 14) | 7 (0, 17) | 8 (3, 13) | 0.53 |
| LVMI (g/m²), median (IQR) | | 74 (58, 90) | 72 (58, 86) | 75 (59, 91) | 0.20 |
| eGFR (mL/min/1.73m²), median (IQR) | | 96 (75, 117) | 102 (80, 124) | 94 (77, 112) | **0.05** |
| ***CARDIOVASCULAR RISK FACTORS AND OUTCOMES*** | | | | | |
| Dyslipidemia, n yes (%) | | 46 (22%) | 12 (15%) | 34 (27%) | 0.56 |

| | | | | | |
|---|---|---|---|---|---|
| Hypertension, n yes (%) | | 42 (20%) | 10 (13%) | 32 (25%) | 0.36 |
| Diabetes, n yes (%) | | 11 (5%) | 1 (1%) | 10 (4%) | 0.21 |
| Smoking status, n (%) | Current smoker | 28 (14%) | 8 (10%) | 20 (16%) | 0.59 |
| | Ex-smoker | 37 (18%) | 9 (11%) | 28 (22%) | |
| | Nonsmoker | 115 (56%) | 38 (48%) | 77 (61%) | |
| | *Missing* | 26 (13%) | 24 (30%) | 2 (2%) | |

**Supplemental Table S4:** Incident stroke/TIA rate in patients with severe or attenuated variants (n=31 patients with unknown variants included in overall count but not analyzed separately).

| | **Total cohort (n = 641)** | **Females** | | | **Males** | | |
|---|---|---|---|---|---|---|---|
| | | **Overall* (n = 414)** | **Attenuated (n = 120)** | **Severe (n = 269)** | **Overall* (n = 227)** | **Attenuated (n = 83)** | **Severe (n = 138)** |
| # Patients ever with stroke/TIA | 57 | 27 | 3 | 22 | 30 | 5 | 24 |
| # Stroke/TIAs | 98 | 40 | 9 | 28 | 58 | 7 | 49 |
| # Patient years | 6308 | 4094 | 1026 | 2913 | 2214 | 625 | 1579 |
| Average # years in study per patient | 9.8 | 9.9 | 8.6 | 10.8 | 9.8 | 7.5 | 11.4 |
| Overall stroke/TIA rate per patient year | 0.016 | 0.0098 | 0.0088 | 0.0096 | 0.026 | 0.011 | 0.031 |
| First stroke/TIA rate per patient year | 0.0090 | 0.0066 | 0.0029 | 0.0076 | 0.014 | 0.0080 | 0.015 |

**Supplemental Table S5:** Incident stroke/TIA rate by GLA and sex across 3 age categories: <40, 40-59 and 60+ years of age.

| **Group** | **Incident stroke/TIA rate** |
|---|---|

| | <40 years old | 40-59 years old | 60+ years old |
|---|---|---|---|
| Male/Severe | 0.0074 | 0.0218 | 0.0317 |
| Male/Attenuated | 0.0000 | 0.0045 | 0.0181 |
| Female/Severe | 0.0027 | 0.0065 | 0.0163 |
| Female/Attenuated | 0.0027 | 0.0027 | 0.0034 |

**Supplemental Table S6:** Incident stroke/TIA rate by sex and cohort (CFDI versus Canadian general population) by 3 age categories: <40, 40-59 and 60+ years of age. **Canadian data doesn't distinguish incident versus recurrent strokes, nor include TIAs.*

| Group | Stroke rate* | | |
|---|---|---|---|
| | <40 years old | 40-59 years old | 60+ years old |
| CFDI Males | 0.0060 | 0.0184 | 0.0231 |
| Canadian Males | 0.0002 | 0.0018 | 0.0085 |
| CFDI Females | 0.0027 | 0.0052 | 0.0139 |
| Canadian Females | 0.0003 | 0.0017 | 0.0089 |

**Supplemental Table S7:** Comparing prescription trends pre- versus post-trial timepoints by GLA severity or sex.

| Prescription pre- versus post-trial | Attenuated GLA (n=50) | Severe GLA (n=137) | p | Female (n=141) | Male (n=52) | p |
|---|---|---|---|---|---|---|
| Stayed on ASA/AP, n (%) | 11 (22%) | 42 (31%) | **0.013** | 40 (28%) | 14 (27%) | 0.60 |
| Deprescribed, n (%) | 16 (32%) | 16 (12) | | 21 (15%) | 11 (21%) | |
| Prescribed, n (%) | 3 (6%) | 13 (9%) | | 15 (11%) | 3 (6%) | |
| Never on ASA or AP, n (%) | 20 (40%) | 66 (48%) | | 65 (46%) | 24 (46%) | |

**Supplemental Table S8:** Characteristics of the 5 patients among the 57 with incident stroke/TIA during the study period who had prior PCI/CABG.

| Sex | GLA Severity | Age at first stroke/ TIA | 10-year ASCVD risk score at year of | Year(s) PCI/CABG | Year(s) stroke/TIA(s) | Lacunar/ small vessel disease suggested? |
|---|---|---|---|---|---|---|

| | | | **first stroke/TIA** | | | |
|---|---|---|---|---|---|---|
| Male | Severe | 37 | 1.15 | 2016 (no MI) | 2008, 2017, 2019 | Yes |
| Male | Severe | 45 | 1.2 | 2014, 2017 (no MI) | 2011, 2015, 2016, 2019 | Yes |
| Female | Severe | 48 | 2.02 | 2010 (no MI) | 2012, 2017 | Yes |
| Male | Severe | 52 | 2.74 | 2015 (no MI) | 2015, 2021 | No |
| Male | Severe | 55 | 3.02 | 2009 (with MI) | 2015 | No |
